# Modeling COVID-19 Aerosol Transmission in Primary Schools

**DOI:** 10.1101/2021.12.08.21267499

**Authors:** Tessa Swanson, Seth Guikema, James Bagian, Claire Payne

## Abstract

Schools must balance public health, education, and social risks associated with returning to in-person learning. These risks are compounded by the ongoing uncertainty about vaccine availability and uptake for children under 12 years of age. In this paper, we show how the risk of infections that result directly from in-class aerosol transmission within an elementary school population can be estimated in order to compare the effects of different countermeasures. We compare the effectiveness of these countermeasures in reducing transmission including required masking at three levels of mask effectiveness, improving room airflow exchange rates, weekly testing of the students, and lunch partitioning. Our results show that multiple layers of interventions are necessary to keep in-class infections relatively low. These results can inform school administrators about how these interventions can help manage COVID-19 spread within their own elementary school populations.

## 1 Introduction

Elementary schools pose a particularly difficult setting for managing risks associated with COVID-19. Children under 12 years of age only became eligible for vaccination in the U.S. in late 2021. Vaccination uptake remains uncertain, and relatively high vaccination rates for this age group will likely not be achieved until well into 2021 in the U.S. In other countries, children remain ineligible for vaccination.

At the same time, elementary schools provide ideal settings for the spread of an aerosol-transmitted virus such as SARS-CoV-2. Students are together in indoor spaces, often in older, poorly ventilated schools for multiple hours per day. In many schools they have shared lunch periods, by necessity without wearing masks while eating, in cafeteria spaces that are often poorly ventilated. Many schools, at least in the U.S., have heating and ventilation systems that are not ideal for reducing the spread of airborne infectious agents such as viruses (Hoover et al., 2021). The degree of filtration of recirculated air is often limited by the ventilations system design and existing equipment, there are strong limitations on the degree of outside air that can be introduced, and in some cases, there is not a central ventilation system in classrooms (Hoover et al., 2021; Li et al., 2007).

There is little doubt that in-person learning is preferred to on-line learning for educational, social development, and psychological reasons for elementary school age children. However, this raises a risk-risk trade-off in which health risks associated with SARS-Cov-2 spread must be balanced with educational, social, and development risks. Schools also face an increasingly difficult challenge with parents becoming increasingly vocal and critical of decisions made by school leadership, and some possible risk management options imposing large costs on school districts.

In this paper we develop a simulation model to help inform risk management decision making by school leaders. This approach is grounded in risk analysis and focuses on comparing the effects of different interventions such as masking, testing, lunch policies, and airflow improvements on the spread of SARS-CoV-2 in elementary schools. The goal of this model is not to precisely predict the number of COVID cases in a school under a particular set of interventions but to compare the influence of different interventions, and combinations of interventions, on the risk of infection in a school. Our results suggest that a defense in depth approach involving multiple interventions is critical to keeping cases low in elementary schools with populations of students that are largely unvaccinated and at the same time providing robustness and resilience in light of the continual mutation of the SARS-CoV-2 virus that can present different risk of infection and disease severity.

## 2 Background

The dominance of aerosol transmission of SARS-CoV-2 means that indoor spaces present increased risk for exposure to the virus, particularly when activities include high respiratory activity such as speaking and eating (Anderson et al., 2020; Asadi et al., 2020; Morawska & Cao, 2020). Simulation modeling offers a way to model these spaces and experiment with different interventions to understand their effects on exposures and infections. With simulation modeling, we can develop digital representations of real-world systems, e.g, a “digital twin,” and carry out experiments that are not otherwise feasible due to scale, cost, or other constraints. Efforts to simulate human interactions and the virus exposures surrounding those interactions provide valuable insights into transmission pathways and possible defenses against transmission. Many advances have been made in simulating aerosol dynamics in indoor environments including restaurants, airplanes, elder care facilities, hospitals, universities, and schools (Azimi & Stephens, 2013; Bazant & Bush, 2021; Curtius et al., 2021; Farthing & Lanzas, 2021; Gröndahl et al., 2021; Jimenez, n.d.; Ranoa et al., 2021).

Given the role of school openings in student and community COVID-19 incidence (Auger et al., 2020; Stein-Zamir et al., 2020) a number of simulation models have been developed to model COVID-19 spread within elementary schools and their surrounding communities, though these models are almost exclusively compartmental SEIR (susceptible, exposed, infected, recovered) models. (Tupper & Colijn, 2021) modeled effects of testing and cohorting to find that interventions triggered by positive tests from symptomatic individuals are relatively ineffective in mitigating outbreaks. (Zhang et al., 2021) modeled a single well-mixed population of students to conclude that without testing and masking, 70%-90% of students would be infected but interventions could reduce student absence days by 80%. (McGee et al., 2021) examined cohorting, surveillance testing through an SEIR model coupled with a social network model to find that cohorting and testing together can substantially reduce cases. Other simulations expand on in-class transmissions to model the impacts of school reopening strategies on not only students but also their households and surrounding communities. (Phillips et al., 2021) used agent-based modeling to model a childcare center and a primary school as well as their respective students’ households to find that reducing class sizes and clustering family members together significantly reduce transmission. (Mele et al., 2021) uses an agent based SEIR model for the state of North Carolina to show that masking in school settings can reduce secondary infections by 23-36% for fully-open schools, accounting for mask quality and fit. (Cohen et al., 2020) created an agent-based model to evaluate several hybrid reopening strategies by primary, middle, and high schools and highlights tradeoffs in terms of total infections versus the number of school days spent at home. (Di Domenico et al., 2021) simulated impacts of school reopenings across all of France based on partial, progressive, or full reopening strategies and found that ICU’s would ultimately become overwhelmed if all schools reopened fully at the same time. (Panovska-Griffiths et al., 2020) also modeled partial versus full reopening along with different testing policies in the UK to find that reopening schools must be accompanied by population-wide testing, contact tracing, and isolation to avoid a consequent national wave of infections.

In contrast to these aforementioned simulation models, we utilize a risk analysis framework to guide our simulation modeling. Risk analysis provides an additional lens for understanding these results and designing future analyses and policies. Risk analysis is a framework concerned with characterizing future scenarios, the consequences of those scenarios, the severity of those consequences, and the uncertainty surrounding all of these components (Aven et al., 2018). In the case of COVID-19 aerosol transmission in elementary schools, infection occurs with different likelihoods in different individuals based on likelihood of exposure and dose response to given exposures. When in a room with an infected individual, exposure is influenced by room dimensions, viral parameters (such as viral load in a droplet, deactivation rate, and settling rate), air change rate in the room, the rate of exhalation from all infected individuals in a room, and the rate of inhalation of susceptible individuals (Anderson et al., 2020; Curtius et al., 2021; Evans, 2020). Dose response modeling is a method for converting exposure to a probability of infection (Watanabe et al., 2010). The likelihood of infection given an exposure concentration for Coronavirus can be modeled by an exponential dose response function (Dabisch et al., 2021; Watanabe et al., 2010). Interventions can be applied to any of these mathematical models, from reducing aerosol exhalation or inhalation with masks, changing the effective airflow rate of a room, removing infected students from rooms through testing and quarantine protocols, or reducing the probability of infection given an exposure concentration by a factor provided by vaccination protections. Defense in depth describes the risk mitigation strategy of layering these interventions to minimize the probability of a failure, in this case infection (Larouzée & Guarnieri, 2015; Sorensen et al., 1999). Of particular note, when integrated into simulation modeling, a defense in depth approach can capture the cumulative effects of multiple interventions to compare not only to a do-nothing scenario, but also to single intervention scenarios. This provides information that enables strategies and tactics to be employed that result in more robust solutions that are less sensitive to the COVID virus variants that will continue to present themselves.

Our model stands out from other simulations modeling COVID-19 spread in elementary schools as we directly estimate SARS-CoV-2 exposure in classrooms and estimate likelihood of infection given that exposure with a dose-response function. Further, we trace students and teachers through their classrooms and lunch periods rather than assuming a network or probability of interactions. This approach captures actual classroom interactions calibrated to the rooms they happen in rather than applying a value of probability of transmission, whether assumed or calculated, that SEIR models rely on. This allows for explicit modeling and comparison of room-level interventions (e.g. HEPA filtering, improving airflow) and individual-level interventions (e.g. masking, testing). Our modeling approach also treats schools as independent entities, enabling the ability to include regional parameters reflecting community disease prevalence, individual student and teacher schedules, and local policy constraints to fit their specific needs. We incorporate defense in depth by considering all combinations of weekly testing, masking, airflow, and lunch policies to evaluate cumulative benefits of these interventions for benefit cost analysis. Finally, we incorporate uncertainty by presenting the distribution of outcomes across all simulation replications and including results bounded by low transmissibility and high transmissibility scenarios to capture uncertainty in the dose-response function which is necessary for reasons such as the presence of COVID variants. With these capabilities, administrators from individual schools or an entire district can adapt this model to determine how to best mitigate risk for students and staff, their families, and the surrounding community.

## 3 Methods

### 3.1 Model Description

This model simulates cumulative exposure of students and teachers in individual class meetings over the course of each day in a semester. The first step of modeling classroom exposure relies heavily on computations developed by (Evans, 2020). We assume air in a classroom is well mixed over the course of a class period and estimate the steady state room concentration of COVID-19 aerosols in viral copies per liter (ρ_*A*_) based on equation (1):

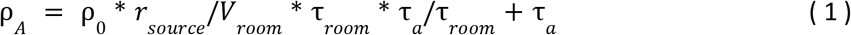

where ρ_0_ is the viral concentration in saliva, *r*_*sorurce*_ is the emission rate of infectious aerosols, *V*_*room*_ is the volume of the room, τ_*room*_ is the air cycle time in a room, and τ_*a*_ is the time for aerosol concentration decay. With this formula as a baseline, we can add or change terms to represent interventions such as masking or air exchange rate changes. We can multiply *r*_*sorurce*_ by the number of infectious people in a room that will be emitting infectious aerosols; we can vary *r*_*sorurce*_ by activity to represent different emission rates of sitting silently, lecturing, or eating; we can introduce masking by reducing *r*_*room*_by a specified factor representing mask effectiveness; we can adjust the τ_*room*_ air change rate to represent interventions that promote air circulation; we can add more decay parameters to represent other interventions that affect the activity of infectious aerosols. Finally, we can convert this room concentration to each class member’s exposure, *N*_*A*_, using equation (2):

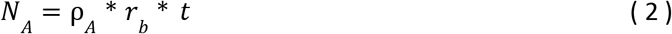

where *r*_*b*_ represents breathing rate and*t* is the amount of time in that room. Again, masking can be implemented in this formula by reducing *r*_*b*_ by a specified factor. The parameter values that we kept constant for calculating room concentration and exposure come from (Evans, 2020) and are summarized in Table 1.

**Table 1:** Constant model parameters

| Parameter | Value |
| --- | --- |
| viral particle settling time | 20 min |
| viral deactivation time | 90 min |
| viral load in saliva | 1000 /nL |
| breathing rate | 10 L/min |
| Student aerosol source rate | 1 nL/min |
| Teaching aerosol source rate | 5 nL/min |
| Eating aerosol source rate | 4 nL/min |

We simulate all class meetings and other activities, such as recess and lunch, over the course of a day and to sum the daily exposure for susceptible (i.e. not previously infected nor vaccinated) individuals. From each individual’s daily exposure *d*, we calculate their probability of infection *P*(*I*) using a dose response function following the exponential distribution shown in equation (3):

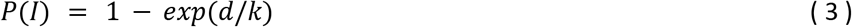

Given the novelty of SARS-CoV-2, the *k* parameter is still unknown, but based on available research we can bound *k* between values and present model results for both of those values (k=75 for high transmissibility case and k=500 for low transmissibility) to communicate this uncertainty and the range of possible outcomes between them (Dabisch et al., 2021; Watanabe et al., 2010). Using each individual’s calculated daily probability of infection, we randomly assign them an infection status based on a Bernoulli distribution. For each infected student, we assign that student to be either symptomatic or asymptomatic again based on a Bernoulli distribution with a specified probability representing an asymptomatic rate of 40% (Tupper & Colijn, 2021). We calculate a period of infectiousness that includes a 2-day latent period and then varies based on whether the student is symptomatic or not, assuming that symptomatic students will stay home after showing symptoms. This period of infectiousness calculation can be manipulated to include weekly testing policies, which may shorten an individual’s period of infectiousness if they stay home following a positive test result.

We inject some infections from outside classrooms at a rate of 0.001% per day. For an elementary school of 500 students, this translates to about one new case every two days. This outside infection rate was typical for many U.S. communities at the time of modeling and can be changed to reflect local community prevalence. We also can randomly assign immunity (from prior infection or vaccination) at the beginning of the semester, in this case only assigning immunity to teachers with a probability of 95% per individual to represent a case where, as in the U.S. as of the fall 2021 semester, students in elementary school are not eligible for vaccination. The remainder of the model involves repeating this process for assigning infections over the course of a semester as we track the previously infected, infectious, and susceptible individuals that will populate classrooms every day. We simulate a semester 1000 times to create probability distributions representing the fraction of students infected over the course of a semester. More details of the algorithms behind this simulation model are available in (Swanson et al., 2021).

### 3.2 Elementary School Data

This model represents a prototypical elementary school with 6 grades. Each grade consists of 3 classes of 25 students and 1 teacher each. This totals to 18 total classes, 450 students, and 18 teachers. Each class has one 60 minute meeting and one 90 minute meeting in the morning separated by a period outside, followed by a 40 minute lunch, and finally a one 50 minute meeting and one 100 minute meeting in the afternoon again separated by outside recess. Each class remains in the same classroom for each class period throughout the day. For lunch, the teachers eat unmasked in the teacher’s lounge. We model three different lunch scenarios for the students: one with all students eating unmasked in the cafeteria with teachers eating together unmasked in a separate lounge, one with each class eating unmasked in their respective classrooms with their teachers, and one with all grades eating unmasked outside with their teachers. We assume all students are unvaccinated and teachers each have a probability of 95% of being fully vaccinated. These immunity rates are input values into the model and so can be adjusted to reflect local vaccination rates for teachers and vaccine rollout for children 5-11 years old.

### 3.3 Scenarios

In order to evaluate the benefits of different interventions, we use the simulation model to run scenarios including different combinations of masking effectiveness, testing policies, lunch policies, and airflow improvements. We assume all students and teachers wear masks, but we vary the effectiveness of masks to compare 40%, 60%, and 95% effectiveness. We evaluate a weekly testing policy by randomly assigning all students a weekday test day and, assuming 100% compliance and 100% test accuracy, remove infectious students from classrooms for the remainder of their infectious period one day after their test day representing a one day turnaround time for receiving test results. This assumption of perfect testing is optimistic and thus an upper bound (best case) on the value of testing. We also test different lunch policies, described above. Finally, we model the effects of airflow improvements, increasing effective air changes per hour (ACH) from a baseline of 4 to a standard of 7 that can be implemented by opening windows or installing HEPA filters.

### 3.4 Assumptions

As with all models, ours includes a number of assumptions and simplifications. We described these assumptions in the model and scenario descriptions and we summarize them in Table 2.

**Table 2:** Model assumptions

| Stage | Assumption |
| --- | --- |
| Exposure | Does not include droplets or fomites given masks effectively protect from droplet borne spread and fomites are not |
|  | considered a significant transmission pathway at this time |
|  | Rooms are well-mixed |
|  | Masks are not worn during lunch |
|  | Transmission does not occur outdoors |
| Outside infections | Occur randomly at a rate of 0.001% per day |
| Period of infectiousness | There is a 2-day lag between getting infected and becoming infectious and/or experiencing symptoms |
|  | 40% of infections are asymptomatic |
|  | Without symptoms or a positive test result, asymptomatic students continue to go to class and are infectious for 1 week |
|  | Symptomatic students stop going to class for the remainder of their infectious period after 1 day of infectiousness |
| Testing (if applicable) | Students are randomly assigned a weekday on which they get tested every week with 100% compliance |
|  | There is a 1 day turnaround time for receiving test results |
|  | Asymptomatic students stop going to class for the remainder of their infectious period after 1 day following their test |
|  | Testing is 100% accurate |

## 4 Results

### 4.1 Scenario Model Outputs

We use box and whisker plots to summarize the percent of infected students over 1000 replications of each scenario. The middle line of each of these plots represents the median proportion of students infected over the course of the modeled semester. The box covers the interquartile range, or the middle 50% of replications, from the 25th- to 75th-percentile. The whiskers, or vertical lines, extend to cover the remainder of the data that falls within 1.5 times the interquartile range, with outliers represented as dots outside of the range of the whiskers. The plot below shows the results of all runs where masks were set to be 40% effective at filtering aerosols exhaled by infected students or teachers as well as inhaled by susceptible students and teachers. We segment these plots by lunch arrangement and compare the scenarios on the x-axis describing combinations of ACH level and whether or not there is a weekly testing requirement. Given the uncertainty about the k parameter in the dose response function as discussed in the Background section, we plot results in Figure 1 for both the high transmissibility case (k=75, in red) and low transmissibility case (k=500, in blue).

**Figure 1:**
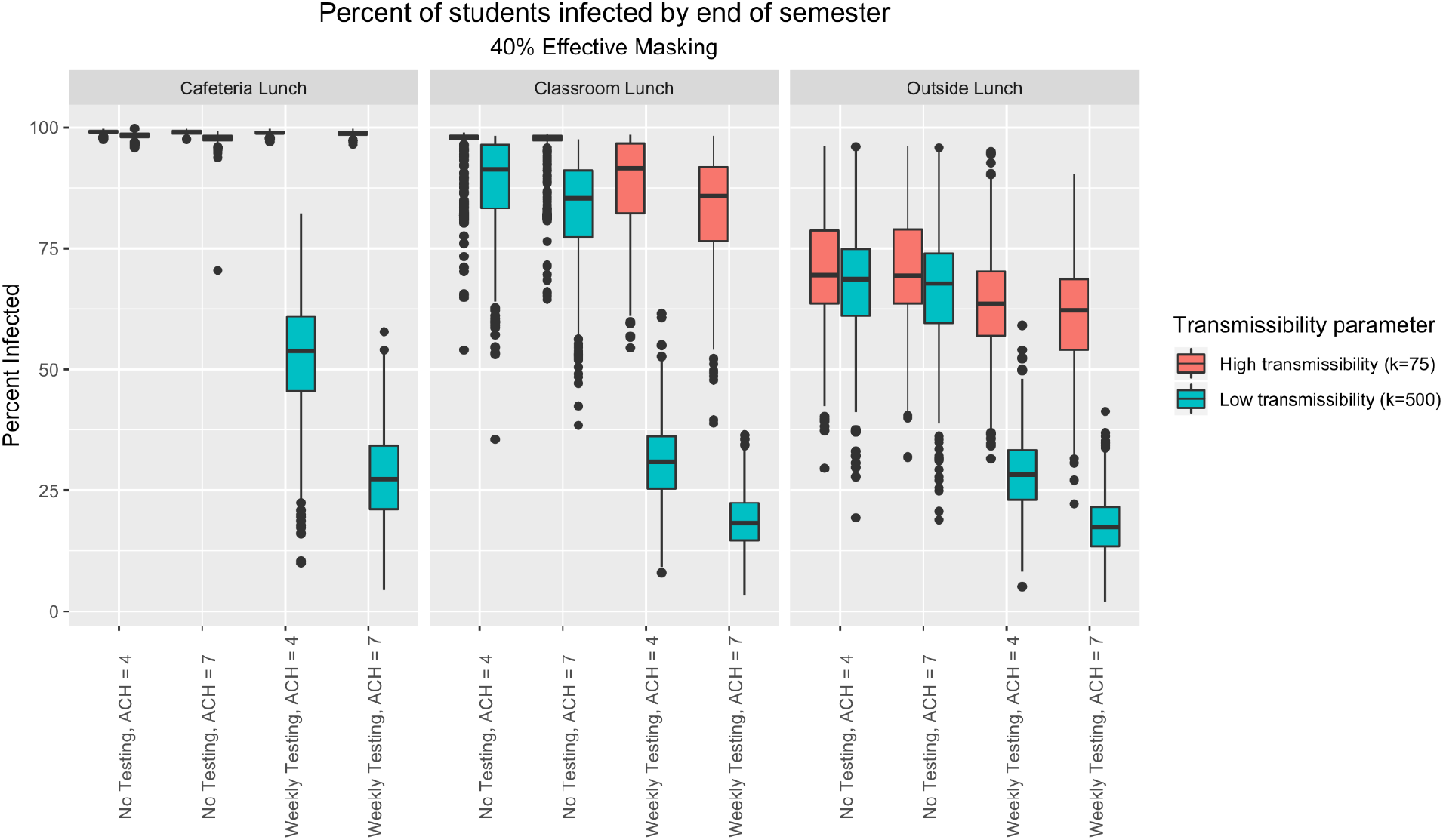
Box and whisker plots comparing scenarios with 40% masking effectiveness

Looking at the results by lunch arrangement, a maskless lunch in the cafeteria leads to nearly all students becoming infected over the course of the semester when there is no testing, with testing reducing infections by about half in the low transmissibility scenario. If the true k value is closer to the high transmissibility case of 75, even testing and subsequent quarantining does not prevent almost all students from becoming infected over the semester consistently across all replications. With a classroom based lunch, the percentage of students infected decreases, but only when testing is enforced does the range decrease below 50% if transmissibility is low. The large distance between the distributions of the low and high transmissibility cases for each of the classroom lunch scenarios reveals high uncertainty that remains even with multiple interventions including implementing testing, raising ACH, and moving lunch to classrooms.

Holding lunch outside leads to a further decrease in expected number of students infected, with testing again leading to a further decrease in the percentage of students infected, more notably in the low transmissibility case. We see across all of these plots that raising the ACH from 4 to 7 does make some difference in decreasing infections, but its effectiveness is more substantial when coupled with weekly testing.

Figure 2 shows these same plots for the 40%, 60%, and 95% mask effectiveness levels.

**Figure 2:**
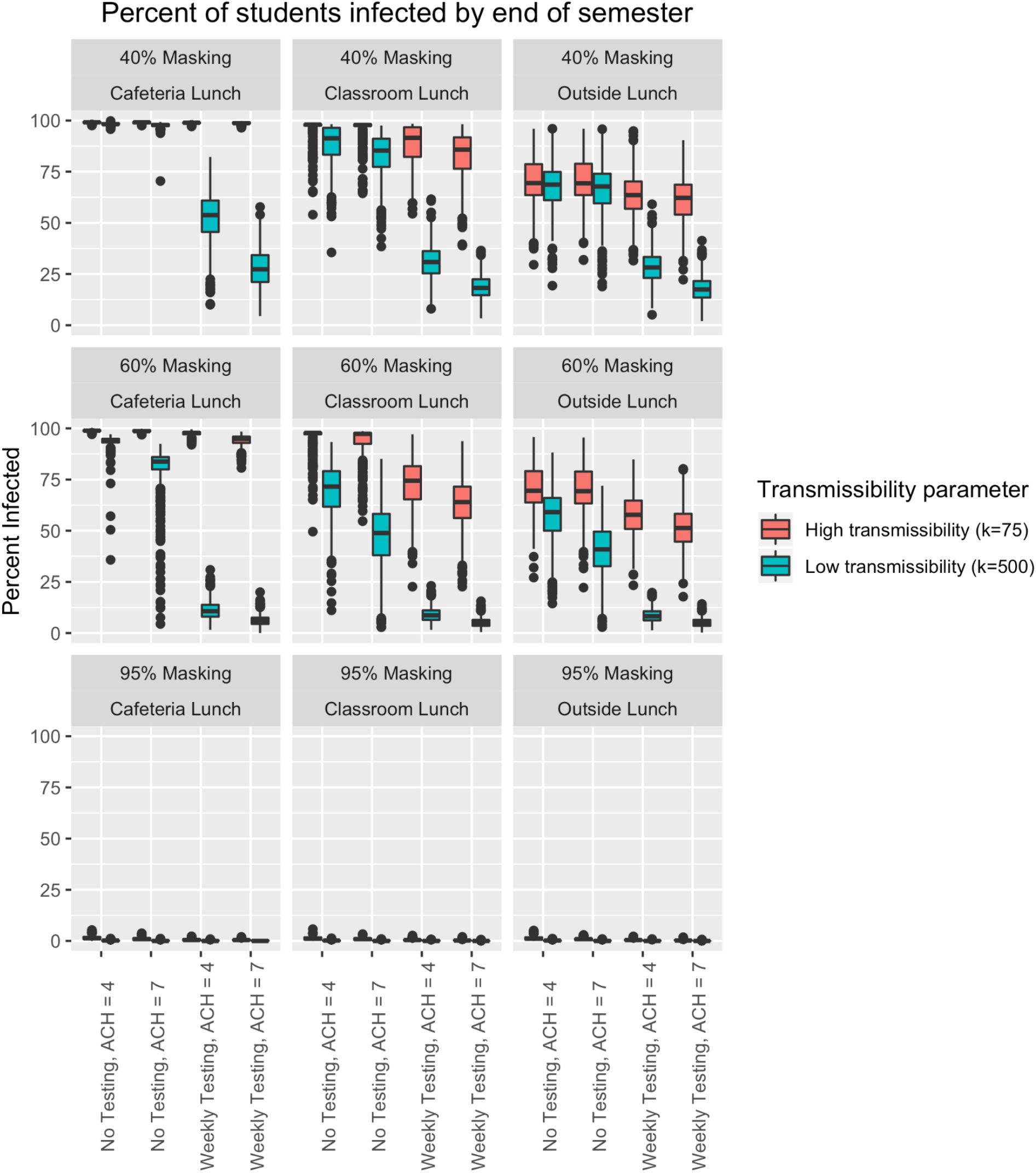
Box and whisker plots comparing scenarios with 40%, 60%, and 95% masking effectiveness

We see similar trends at the 60% effective masking level as the 40% effective masking level, with testing and lunch policy having the greatest impact on reducing the percentage of infected students. At the 95% effective masking level, we see very few student infections over the course of the semester even with no other interventions.

### 4.2 Linear Regression Modeling

To confirm these trends, we performed linear regression analysis on the results. We create a multivariate linear model and summarize the results in Table 3 below. We treat all variables as categorical variables and utilize reference cell coding such that the reference scenario values were high transmissibility, ACH = 4, no weekly testing, lunch in the cafeteria, and 40% effective masking. The response variable in this model is the percent of students infected.

**Table 3:** Multiple linear regression model for estimating percentage of students infected

| Parameter | Coefficient | p-value |
| --- | --- | --- |
| Intercept | 1.06 | <2e-16 |
| Low transmissibility | -0.246 | <2e-16 |
| ACH = 7 | -0.0416 | <2e-16 |
| Weekly testing | -0.222 | <2e-16 |
| Outside lunch | -0.186 | <2e-16 |
| Classroom lunch | -0.0881 | <2e-16 |
| 60% effective masking | -0.126 | <2e-16 |
| 95% effective masking | -0.705 | <2e-16 |
| Residual standard error: 0.1838 on 71992 degrees of freedom<br>Multiple R-squared: 0.7909, Adjusted R-squared: 0.7909<br>F-statistic: 3.891e+04 on 7 and 71992 DF, p-value: < 2.2e-16 |  |  |

All coefficients are statistically significant at the 95% confidence level and, as expected, all interventions have negative coefficients, meaning they reduce the percentage of students infected over the course of a semester. The greater than 1 intercept coefficient indicates all students are expected to be infected under the reference scenario with high transmissibility, while each intervention decreases that proportion of students infected by an average of their respective coefficient values across all scenarios. Other than 95% effective masking, weekly testing of all students and teachers has the largest effect on risk of infection, followed by moving lunch outside.

In order to capture the interactive dynamics of these interventions, we also generate a multiple linear regression model with interactions. We summarize the coefficients of these variables and their significance in Table 4. A negative coefficient for the interaction term indicates those two interventions together offer synergistic effects and decrease the percentage of infected students even more than those two interventions independently.

**Table 4:** Multiple linear regression with interactions model for estimating percentage of students infected

| Parameter | Coefficient | p-value |
| --- | --- | --- |
| Intercept | 1.085201 | <2e-16 |
| Low transmissibility | -0.134315 | <2e-16 |
| ACH = 7 | -0.018723 | <2e-16 |
| Weekly testing | -0.155381 | <2e-16 |
| Outside lunch | -0.351093 | <2e-16 |
| Classroom lunch | -0.087509 | <2e-16 |
| 60% effective masking | 0.003439 | 0.121762 |
| 95% effective masking | -1.149467 | <2e-16 |
| Low transmissibility *<br>ACH = 7 | -0.048766 | <2e-16 |
| Low transmissibility *<br>Weekly testing | -0.301809 | <2e-16 |
| Low transmissibility *<br>Outside lunch | 0.079506 | <2e-16 |
| Low transmissibility *<br>Classroom lunch | -0.016352 | <2e-16 |
| Low transmissibility *<br>60% effective masking | -0.153632 | <2e-16 |
| Low transmissibility *<br>95% effective masking | 0.281586 | <2e-16 |
| ACH = 7 *<br>Weekly testing | -0.013865 | <2e-16 |
| ACH = 7 *<br>Outside lunch | 0.005324 | 0.003344 |
| ACH = 7 *<br>Classroom lunch | -0.010338 | 1.22e-08 |
| ACH = 7 *<br>60% effective masking | -0.019826 | <2e-16 |
| ACH = 7 *<br>95% effective masking | 0.050056 | <2e-16 |
| Weekly testing *<br>Outside lunch | 0.050740 | <2e-16 |
| Weekly testing *<br>Classroom lunch | -0.014467 | 1.56e-15 |
| Weekly testing *<br>60% effective masking | -0.060099 | <2e-16 |
| Weekly testing *<br>95% effective masking | 0.297219 | <2e-16 |
| Outside lunch *<br>60% effective masking | 0.008589 | 0.000111 |
| Outside lunch *<br>95% effective masking | 0.282852 | <2e-16 |
| Classroom lunch *<br>60% effective masking | -0.047947 | <2e-16 |
| Classroom lunch *<br>95% effective masking | 0.107634 | <2e-16 |
| Residual standard error: 0.09938 on 71973 degrees of freedom<br>Multiple R-squared: 0.9389, Adjusted R-squared: 0.9389<br>F-statistic: 4.255e+04 on 26 and 71973 DF, p-value: < 2.2e-16 |  |  |

From these results, we see that only outside lunch and 95% effective masking have positive interactive terms, indicating the added benefit of other interventions are not as high as the net benefit of those interventions on their own, but still offer additional benefits than either of the interventions alone. All other options appear to offer compounding effects at the first-order interactive level, meaning their benefits to reducing the percentage of students infected are even greater together than just adding up their individual benefits.

We also summarize the interactive effects of these interventions in Figure 3 to highlight where interventions have amplified impacts when coupled with other interventions. We model the high and low transmissibility case separately for these plots and we remove the 95% masking variable, as that intervention alone can reduce the percentage of student infections to nearly zero. The value represented by a shaded square at the intersection of two variables is equal to the cumulative impact of combining those interventions by summing their individual coefficients as well as their interaction term.

**Figure 3:**
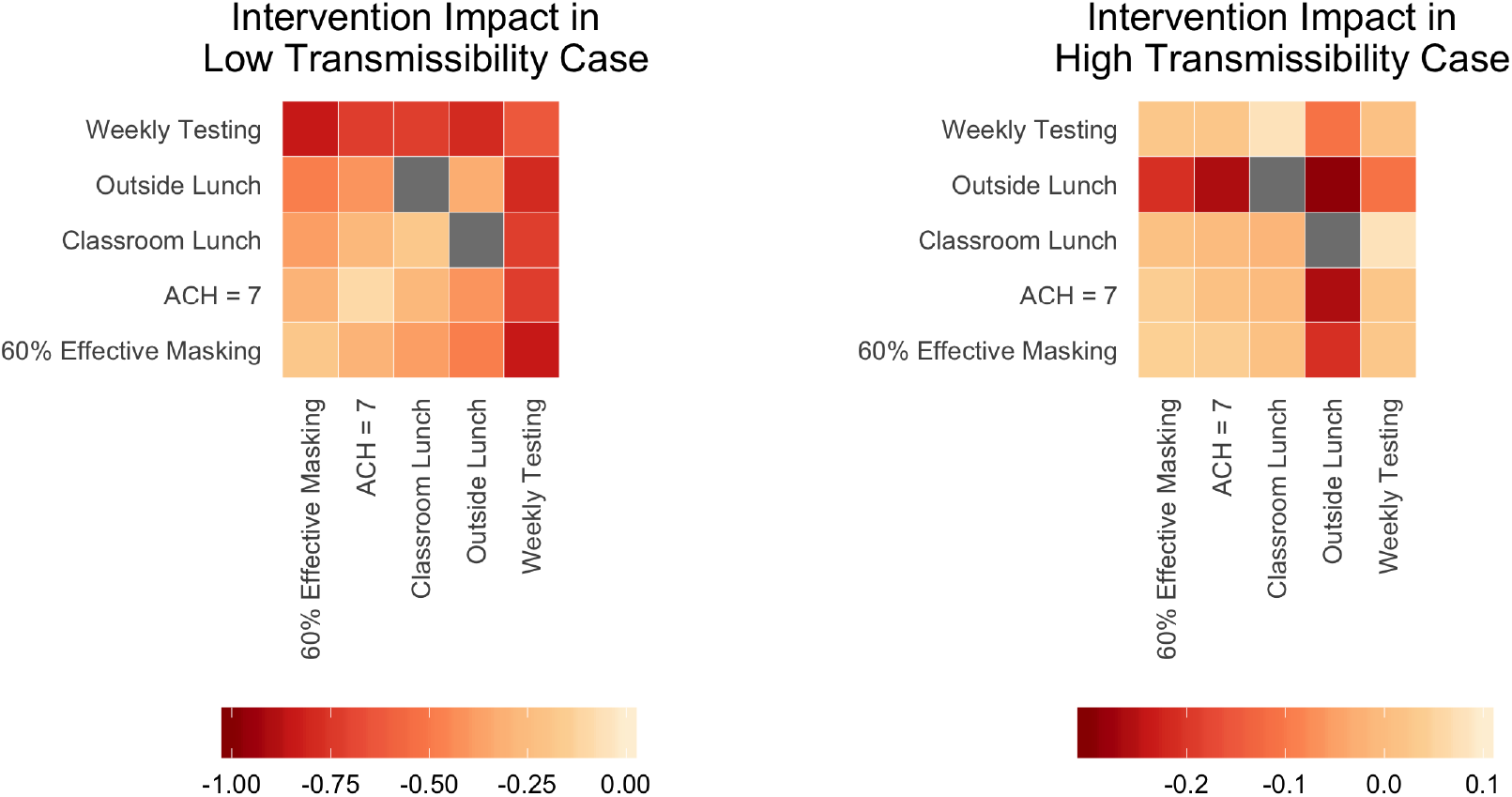
Cumulative impact of paired interventions by transmissibility case

The darker red values indicate stronger interactive effects. In the low transmissibility case, weekly testing offers the highest additional impact of all interventions. In the high transmissibility case, moving lunch outside offers the highest additional impact of all interventions.

## 5 Discussion

We present a number of scenarios showing the compounding effects of various interventions available for elementary schools. We demonstrate it takes a defense in depth approach to manage infection risk among unvaccinated elementary school students through our box and whisker plots showing the spread of simulated outcomes for each combination of interventions and multivariate linear models showing the impact of those interventions alone and in combination with others. Mass maskless lunches are particularly risky, while outdoor lunches present the least amount of risk but may be prohibited by external factors like weather. Effective masking remains critical, and while 95% effectiveness across all students may not be feasible these results support enforcement of proper mask wearing in elementary schools. Testing is also critical, where weekly testing may be effective enough with low to moderate transmission rates but more frequent testing may be necessary in situations of high viral transmission. This simulation model can readily be adapted to include multiple weekly testing days to show if frequent testing may be effective for the higher transmissibility case. Increasing the effective air change rate by opening windows or installing HEPA filters is perhaps the easiest to implement, but does not totally offset the risks of in person lunches or no testing and is most effective in combination with testing. Ultimately, we demonstrate that with added interventions comes a decrease in the percentage of students infected as well as a decrease in the spread of modeled outcomes between high and low transmissibility scenarios. A defense in depth approach is critical for effectively and efficiently managing risk of student infection and the subsequent risks of community spread from elementary schools under the high level of uncertainty surrounding transmission of SARS-CoV-2 for multiple reasons such as new viral variants, community prevalence, and community specific factors.

## 6 Conclusion

Elementary schools face unique risks surrounding in-person learning during the COVID-19 pandemic as unvaccinated students assemble and then disperse back to their households on a nearly daily basis. While many of the mechanisms of transmission remain uncertain, minimizing transmission in primary school environments is critical for reducing risk for students, teachers, their families, and surrounding community members. At the same time, eliminating in person learning completely poses costs on learning outcomes for students and childcare burdens on families, particularly afflicting marginalized populations. Even after vaccinations for 5- to 11-year olds are available, distribution will take time and schools may lack authority, ability, or will to mandate and verify vaccination status. Also, depending on the strain of COVID variant the effectiveness of vaccination or past immunity may be dramatically diminished.

Simulation modeling can aid in minimizing transmission and subsequent risk to families and communities by enabling decision makers to evaluate parameters and interventions specific to the needs of individual schools and their localities. We only show results for a prototypical school building and class schedule, but these results show how interventions like masking, air filtering, testing, and distancing during lunch offer cumulative effects to mitigate risk. In addition to protecting students, results from simulation can encourage greater transparency and more effective communication between school administrators and their parents and teachers, which can foster community buy-in and cooperation for improved outcomes. With such tools at their disposal, school administrators are responsible to their students, teachers, and communities for utilizing all of their resources to effectively manage risk.

## Data Availability

No data from actual schools is used in this paper. The code used in the simulation underlying this paper will be made available to others via a public reponsitory after acceptance of the peer-reviewed journal of this paper.

## Notes

### Competing Interest Statement

The authors have declared no competing interest.

### Funding Statement

This study was funded by internal University of Michigan funds through the University of Michigan College of Engineering COVID skunkworks program.

